# Anaphylactic events in mRNA vaccines: a reporting case-control study

**DOI:** 10.1101/2021.07.19.21260714

**Authors:** Chris von Csefalvay

## Abstract

**Background:** mRNA vaccines are a novel method of eliciting immunity, and play a significant role in the global fight against COVID-19. Anaphylactic reactions are a widespread concern driving vaccine hesitancy due to the serious and potentially fatal nature of anaphylaxis. A quantitative estimation of the risk of anaphylactic and anaphylactoid reactions deriving from mRNA vaccines is of a significant public health importance.

**Objective:** To estimate the relative Reporting Odds Ratio of anaphylactic and anaphylactoid reactions following mRNA vaccination vis-a-vis other vaccinations.

**Design:** Reporting case-control study.

**Setting:** Persons reporting adverse events following vaccination to VAERS whose reports were received between 01 January 2000 and 02 July 2021, inclusive.

**Patients:** Each case of anaphylaxis or anaphylactoid reaction was matched with 2.7 unique controls on average, by gender and age rounded to the nearest integer.

**Measurements:** Overall and stratified Reporting Odds Ratios (ROR) were calculated. Stratified contingency tables were tested for homogeneity using the Breslow-Day procedure, and Cochran-Mantel-Haenszel statistics were calculated to test the hypothesis of a ROR of unity.

**Results:** 2,665 cases of anaphylaxis or anaphylactoid reactions and 7,125 controls of non-anaphylactic/anaphylactoid reports were compared. The ROR of an anaphylactic or anaphylactoid reaction was 1.325 (95% CI: 1.212 – 1.448, *p* < 0.001). The matched set of cases and controls revealed an expected inhomogeneity by sex (with women slightly more likely to report anaphylactic presentations) and age band strata (with a bimodal distribution that reflects the common incidence of anaphylactic and allergic pathologies). No significant increase in the risk of anaphylactic adverse events was witnessed among persons who self-reported previous allergic reactions to vaccines. A slightly elevated ROR was observed with patients who reported a history of allergic reactions to NSAIDs and/or fluoroquinolone antibiotics. The precise meaning and relevance of this finding remains to be elucidated.

**Limitations:** As a reporting study using data from VAERS, our analysis is subject tunder- and overreporting, the extent of each of which is not known with any degree of precision. Since the Emergency Use Authorizations for both mRNA vaccines mandate reporting of all serious adverse events, reporting bias is likely in favour of non-mRNA vaccines, where such reporting is not mandatory in adults. Consequently, this analysis may exaggerate the ROR of anaphylactic and anaphylactoid events associated with mRNA vaccines, which may in reality be significantly lower.

**Conclusions:** mRNA vaccination is not associated with a statistically significant higher risk of reporting an anaphylactic adverse event to VAERS. Anaphylaxis is a serious but very rare complication of all immunisations. No significant increase in reporting odds was found in any age group or gender, nor in most cases of previously known allergic adverse events in relation to vaccines. This study contributes to the growing body of evidence proving the safety and tolerability of mRNA vaccines.

## 1 Introduction

### 1.1 Background

Anaphylaxis describes a severe systemic allergic reaction to an antigen, resulting in large-scale mast cell degranulation and, consequently, histamine release. ^[1]^ In clinical practice, this typically manifests as an acute onset of respiratory symptoms, bronchoconstriction, urticaria, flushing, gastrointestinal symptoms (nausea, vomiting, diarrhoea) and hypotension tending towards circulatory collapse.^[2]^ Anaphylactic reactions to vaccines, while fortunately vanishingly rare, are well-documented in relation to a wide range of vaccines.^[3–6]^ The mRNA-based vaccines approved in the United States (BNT162b2/tozinameran, commonly known as the Pfizer/BioNTech vaccine, and mRNA-1273/elasomeran, commonly known as the Moderna vaccine) are no exceptions to this phenomenon. Rare episodes of anaphylaxis have been documented in the context of both the Moderna^[7]^ and the Pfizer/BioNTech vaccine.^[8]^

Because of the severity and, absent rapid and appropriate medical intervention, life-threatening nature of anaphylactic reactions, severe allergic adverse events have been key drivers behind vaccine hesitancy.^[9–11]^ Since the Pfizer/BioNTech and Moderna vaccines represent the first prophylactic mRNA based vaccines in wider public health use, understanding the true risk of anaphylaxis from these novel vaccines is crucial to address vaccine hesitancy and safeguard the goals of the COVID-19 vaccination programmes world-wide.

### 1.2 Objectives

The objective of this study was threefold:

1. to investigate the effect of mRNA-based versus non-mRNA-based vaccinations on the likelihood of reporting an anaphylactic symptom to VAERS as opposed to a non-anaphylactic symptom,
2. to quantify whether this effect is homogenous across strata of age groups and sex, and
3. to identify whether certain self-reported allergies are associated with a higher likelihood of reporting an anaphylactic symptom to VAERS as opposed to a non-anaphylactic symptom.

It was hypothesised at the outset that the mRNA-based vaccines would not be associated with a statistically significant elevated ROR (*ROR* > 3.0 at *p* < 0.05) in the whole-population samples. In addition, we hypothesised with respect to the breakdown by sex that while women would be slightly more likely to present with anaphylactic events due to the higher prevalence of anaphylaxis in women in general,^[12]^ the relative difference in odds ratios would be quite modest. We further hypothesised that age strata would also yield a statistically significant inhomogeneity, but that the effect, here, too, would be quite low.

## 2 Methods

### 2.1 Study design

A case-control study, adhering to the STROBE Statement’s methodology and best practices, ^[13]^ was performed on reports to VAERS received between 01 January 2000 and 02 July 2021. Reports were excluded if they did not name a specific vaccine type (VAERS parameter `VAX_TYPE` of `UNK`), or if they pertained to an unidentified COVID-19 vaccine (VAERS parameter `VAX_NAME` of `COVID19 (COVID19 (UNKNOWN))`). Cases of anaphylaxis were identified by the VAERS symptom description, and mRNA vaccines by the name of the specific vaccine in respect of which the report was made. These were matched against controls from the same subset of VAERS reports that reported non-anaphylactic reactions, with matching by age (rounded to the nearest integer) and reported gender.

Comparative statistical analyses were then utilised to determine statistical associations of risk. In particular, stratified odds ratios were calculated for both the crude and the age- and gender-matched sets of reports by gender, age band and reported pre-existing hypersensitivities. For stratified tables, the Breslow-Day statistic was used to test for homogeneity *inter strata* and Cochran-Mantel-Haenszel statistics were used to test for the hypothesis that the pooled odds ratios equal unity. Where odds ratios were calculated per strata, Fisher’s exact test was used to determine independence of variables.

### 2.2 Data sources

Data on adverse events following immunisations was obtained from VAERS via vaers.hhs.gov on 12 July 2021. The data files from 2000 onwards were loaded and processed using Python 3.7.5 (Python Software Foundation, Fredericksburg, VA) and version 1.3.0 of the `pandas` package.^[14]^ Ingested raw data thus included reports received by VAERS between 01 January 2000 and 02 July 2021. Altogether, 1,018,240 reports were ingested in respect of the period under examination.

Of these, we have excluded 140,270 reports for a failure to state a valid age and 18,974 reports for specifying no or unknown sex. In addition, 4,660 reports were excluded for disclosing an unknown vaccine type (`VAX_TYPE` VAERS attribute of value `UNK`) and 744 reports were excluded for documenting the administration of an unknown COVID-19 vaccine (`VAX_NAME` VAERS attribute of `COVID19 (COVID19 (UNKNOWN))`). Following these exclusions, 853,592 eligible reports were processed.

A flow chart of this process is presented as Figure 1.

**Figure 1:**
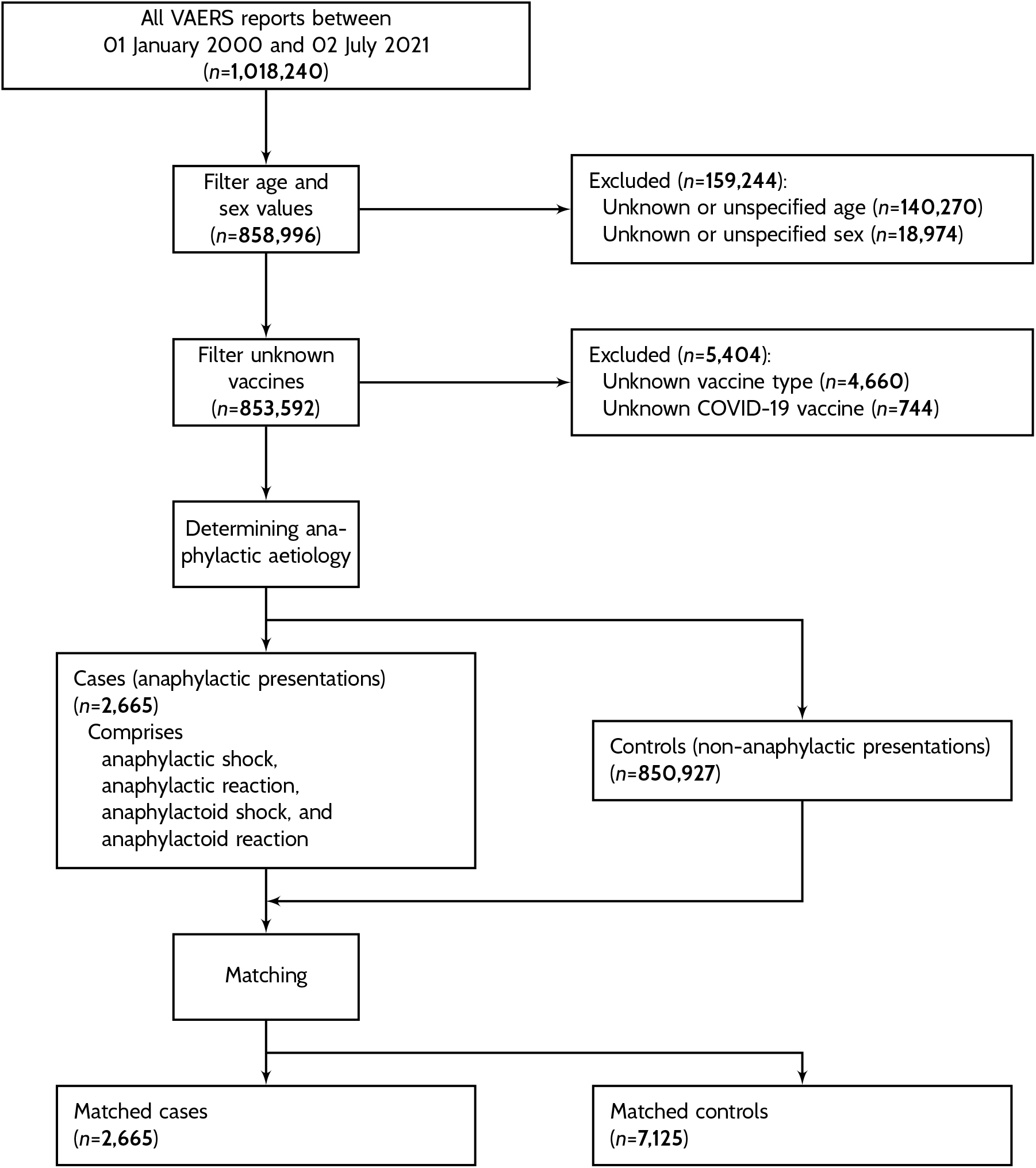
Patient disposition flow chart.

### 2.3 Identifying cases

Cases were defined as reports to VAERS that reported one of four anaphylactic conditions. These were selected from the Preferred Terms (PTs) that are classified by the MedDRA High Level Term (HLT) of Anaphylactic and anaphylactoid responses (MedDRA ID 10077535), under the explicit exclusion of three PTs that have no relevance for vaccine administration: dialysis membrane reactions (MedDRA ID 10076665), anaphylactic transfusion reactions (MedDRA ID 10067113) and anaphylactoid syndrome of pregnancy (MedDRA ID 10067010). The ontology of the included diagnostic categories is presented in Table 1.

**Table 1:** MedDRA ontology of included diagnostic categories. Preferred Terms that are considered an anaphylactic reaction are set in bold type.

| SOC | HLGT | HLT | PT |
| --- | --- | --- | --- |
| Immune system disorders<br>10021428 | Allergic conditions<br>10001708 | Anaphylactic and<br>anaphylactoid<br>responses<br>10077535 | **Anaphylactic reaction**<br>**10002198** |
|  |  |  | **Anaphylactic shock**<br>**10002199** |
|  |  |  | **Anaphylactoid reaction**<br>**10002216** |
|  |  |  | **Anaphylactoid shock**<br>**10063119** |
|  |  |  | Dialysis membrane reaction<br>10076665 |
|  |  |  | Anaphylactic transfusion reaction<br>10067113 |
|  |  |  | Anaphylactoid syndrome<br>of pregnancy<br>10067010 |

While MedDRA offers a Standardised MedDRA Query (SMQ) corresponding to anaphylaxis, this includes a wide range of clinical entities. Some would introduce unnecessary confounders (such as hereditary disorders presenting with anaphylactic symptoms or angioedema, e.g. C1 esterase inhibitor deficiency), while others would lay the analysis open to clearly unconnected events (such as transfusion related anaphylactic events). For this reason, we chose to construe anaphylaxis narrowly. This yielded 2,665 cases of anaphylactic or anaphylactoid symptoms.

### 2.4 Identifying controls

We defined the possible pool of controls as any eligible reports that did not disclose an anaphylactic presentation. 850,927 eligible reports constituted the pool of potential controls.

### 2.5 Identifying exposure

Exposure was defined as the receipt of an mRNA vaccine, regardless of the order of administration. A report was deemed to pertain to an mRNA vaccine if the vaccine administered had a `VAX_NAME` VAERS attribute of

- `COVID19 (COVID19 (MODERNA))` (elasomeran), or
- `COVID19 (COVID19 (PFIZER-BIONTECH))` (tozinameran).

### 2.6 Case-control matching

In order to reduce the confounding impacts of sex (due to the greater propensity of women to develop anaphylactic presentations) and age (due to the unusual age profile of the COVID-19 vaccinated demographic owing to the focus of early vaccination campaigns on the elderly and vulnerable), matching was performed over age (rounded to the nearest integer) and sex. Matching was performed over the exact age, even if age banding was used later for stratification, so as to avoid ‘edge effects’ where inhomogenous distributions of cases within a stratum, peaking towards one or both of the margins, create additional confounding. Separability was relatively weak but present at 55.51% accuracy over 100 balanced models, suggesting that the benefits of matching might be modest but perceptible. The 2,665 cases were matched to 7,125 controls using the propensity matching functionality of `pymatch`. This gives an overall case:control ratio of 2.673 controls per case. The characteristics of the cases and the controls after matching are presented in Table 2.

**Table 2:** Descriptive statistics and exposure data of cases and controls.

|  | Cases<br>(n = 2,665) | Controls<br>(n = 7,125) |
| --- | --- | --- |
| Sex |  |  |
| Male | 732 (28%) | 1,788 (25%) |
| Female | 1,933 (72%) | 5,337 (75%) |
| Age group |  |  |
| <18 | 596 (22%) | 1,549 (22%) |
| 19-25 | 185 (7%) | 448 (6%) |
| 26-35 | 368 (14%) | 924 (13%) |
| 36-45 | 483 (18%) | 1,267 (18%) |
| 46-55 | 433 (16%) | 1,147 (16%) |
| 56-65 | 326 (12%) | 955 (13%) |
| 66-75 | 196 (7%) | 538 (8%) |
| 76-85 | 68 (3%) | 231 (3%) |
| 86-95 | 7 (<1%) | 61 (1%) |
| >95 | 0 (0%) | 3 (<1%) |
| mRNA vaccine exposure |  |  |
| Yes | 1,352 (51%) | 3,116 (56%) |
| No | 1,313 (49%) | 4,009 (44%) |

At an assumed 40% exposure rate among controls, a power of 0.95 at a 95% two-sided confidence level for an odds ratio at or above 1.5 would require between 437 and 453 cases and 1,168 to 1,211 controls, depending on whether Kelsey’s or Fleiss’s corrected method is used. ^[15,16]^ At almost six times the required value, this case-control ratio gives a fairly high predictive power for odds ratios at least as extreme as 1.5.

### 2.7 Statistical analysis

For the purposes of stratification (but not matching), ages were divided into age bands, with a separate age band for persons under the age of 18, 18-25 and thereafter in ten-year increments until the age of 95. A residual age band for persons aged 96 and above was constructed. Following this segmentation, the analysis proceeded in three stages.

First, crude odds ratios were calculated for the entire sample before matching, along with pooled odds ratios by sex and age band. For the analysis of stratified contingency tables, the Breslow-Day statistics were used to test for the homogeneity of odds ratios and the Cochran-Mantel-Haenszel test was used to test for a pooled odds ratio of unity. Statistical analysis used the `StratifiedTable` class of `statsmodels` 0.12.2. ^[17]^

In the second stage, the same odds ratios were calculated, using the same stratification, over the age- and gender-matched sample. This step, too, utilised Breslow-Day statistics for homogeneity *inter strata* and the Cochran-Mantel-Haenszel test was used to test for a pooled odds ratio of unity. Where appropriate, i.e. where the Breslow-Day statistics indicated inhomogeneity, odds ratios by stratum were calculated and Fisher’s exact test was performed to identify their statistical significance.

Finally, odds ratios were calculated specifically for previous allergies. Using regular expressions, the `ALLERGIES` field of each VAERS entry was examined for 17 individual classes of allergens. Since this is a free text field, regular expressions had to be designed to capture most common ways of referring to a class of drugs or prominent instances thereof. The list below reproduces, in parentheses, exemplary items that would respond to the class’s defining regular expressions.

- Opioids (e.g. `CDN`, `Oxycontin`, `pentazocine`)
- Latex (e.g. `Latex`)
- Macrolide and aminoglycoside antibiotics (e.g. `azithromycin`, `Z-pack`)
- Tetracycline antibiotics (e.g. `tigecycline`, `doxycycline`)
- Sulfas (e.g. `Bactrim`, `sulfonamides`)
- Beta-lactam antibiotics (e.g. `Augmentin`, `penicillin`, `cefazolin`)
- Fluoroquinolone antibiotics (e.g. `Cipro`, `levofloxacin`)
- Hydrochlorothiazide (e.g. `HCTZ`, `hydrochlorothiazide`)
- Glycols (e.g. `PEG`, `propylene glycol`)
- Local anaesthetics (e.g. `lidocaine`)
- Known vaccine hypersensitivities (e.g. `Hepatitis vaccine`, `Prevnar`)
- Fish and seafood (e.g. `mussels`, `crab`, `seafood`)
- Insect stings (e.g. `bee venom`, `wasps`, `bee strings`)
- Eggs (e.g. `egg whites`, `egg protein`)
- NSAIDs (e.g. `Motrin`, `Aspirin`, `naproxen`)
- Peanuts and tree nuts (e.g. `walnuts`, `peanuts`)

For the statistical analysis of stated allergies, only entries with a valid entry in the `ALLERGY` field were considered, and no crude (unmatched) values were calculated.

## 3 Results

### 3.1 Anaphylactic events and mRNA vaccination

The ROR of anaphylactic events among mRNA vaccination versus other events and non-mRNA vaccines was 1.325 (95% CI: 1.212 – 1.448). This result was statistically significant at *p* < 0.001, but quite close to unity, indicating the absence of a strong statistical association between anaphylactic events and mRNA vaccination. Persons reporting an anaphylactic event are thus, as hypothesised (see Subsection 1.2), not significantly more likely to have received an mRNA vaccine than a non-mRNA vaccine.

### 3.2 Sex and anaphylaxis risk

It was one of our initial hypotheses that the ROR of anaphylactic events after mRNA vaccines would be slightly higher for women due to the overall higher prevalence of anaphylaxis in female populations, but that such difference would not be highly statistically significant (see Subsection 1.2]). This is confirmed by the results of our analysis, but the overall size of the effect is, as predicted, quite low.

The ROR for female recipients (1.462, 95% CI: 1.316 – 1.624) was slightly higher than for male recipients (1.133, 95% CI: 0.937 – 1.370), and unlike for the latter group, it was statistically significant (at *p* < 0.001). The Breslow-Day statistic (*χ*^2^ = 5.314, *p* = 0.021) was statistically significant, allowing us to reject the null hypothesis that the strata have a common odds ratio. As such, the statistical analysis confirms our hypothesis of a higher anaphylactic risk for female recipients, but the overall odds ratios suggest that this difference is quite modest at best. This analysis thus does not evidence a highly significant sex-dependent differential risk of anaphylaxis.

### 3.3 Age and anaphylaxis risk

The Breslow-Day test (*χ*^2^ = 25.934, *p* = 0.001) once again indicates heterogeneity between age bands. As Figure 2 indicates, only five age bands (26 – 35, 46 – 55, 56 – 65, 66 – 76 and 76 – 85) show statistically significant results. In all of these cases, the reporting odds ratio is modestly above unity but below 2.0, the minimum value commonly agreed to be the lowest possible cut-off for signal generation. Taken together, it appears from the data that the risk of anaphylactic reactions bears a statistically significant relationship to the likelihood of having received an mRNA vaccine, but that the difference in likelihood is quite small. The age distribution suggests a weak bimodal distribution, peaking in the 26 – 35 age range and once again, much stronger, for the 56 – 65 and 66 – 75 age groups. This reflects on one hand the commonly evidenced peak age of anaphylaxis in the fourth decade of life, ^[18]^ along with the relatively higher proportion of elderly vaccine recipients with comorbidities that are associated with allergic or anaphylactic presentations.

**Figure 2:**
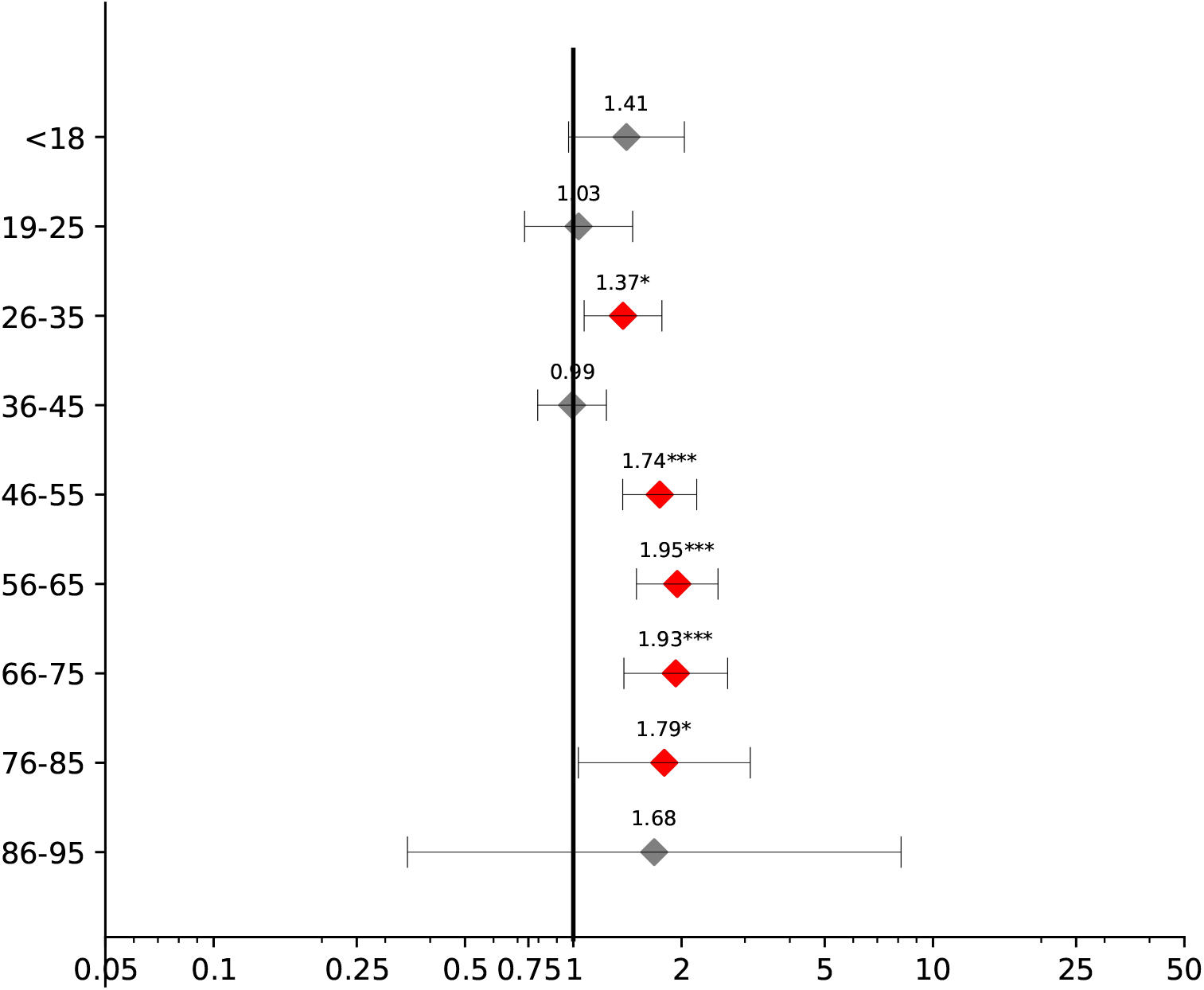
Forest plot of reporting odds ratio of anaphylactic reactions upon receipt of an mRNA vaccine by age band.

Considered as a whole, there are clear age differences in the risk of anaphylaxis, with recipients in the sixth to eighth decade of their lives at highest risk. Nevertheless, this differential increase in risk is very low, and in no age group is there evidence of a strong safety signal. Consequently, no age-dependent safety signals vis-a-vis anaphylaxis emerge from this analysis. As hypothesised, the data confirms mRNA vaccines are not statistically significantly less safe than non-mRNA vaccines, regardless of age group.

### 3.4 Previous allergies and anaphylaxis risk

Of the 17 classes of allergens isolated from the `ALLERGY` free text field, 14 (82.4%) were detected in both the control and case groups. A Breslow-Day statistic of *χ*^2^ = 16.795 at *p* = 0.209 indicates that we cannot reject a null hypothesis of homogeneity. The Cochran-Mantel-Haenszel metric (*χ*^2^ = 9.666, *p* = 0.002) suggests that in the presence of homogeneity, the shared odds ratio is not statistically significantly likely to be at unity. This evidence suggests that while individuals who have previous allergic aetiologies might experience anaphylaxis following mRNA vaccines at a slightly higher rate (pooled ROR: 1.367, 95% CI: 1.122 – 1.666) than the general population (ROR: 1.325, 95% CI: 1.212 – 1.448), no individual allergic predisposition is likely to strongly predispose to anaphylaxis following mRNA vaccination.

This is reflected by the fact that of the 14 relevant categories of allergies, only two were statistically significant. It is notable that these two had also the highest RORs, and the only RORs exceeding 2 (see Figure 3). Pre-existing allergies to NSAIDs (ROR: 3.382, 95% CI: 1.362 – 8.399, *p* < 0.01) and fluoroquinolone antibiotics (ROR: 5.586, 95% CI: 1.119 – 27.900) are associated with higher RORs of anaphylactic reactions. Notably, there is no statistically significant association between pre-existing reports of allergic events (of any severity) from other vaccines, which is in line with the fundamental biochemical and compositional differences between mRNA vaccines and non-mRNA vaccines.

**Figure 3:**
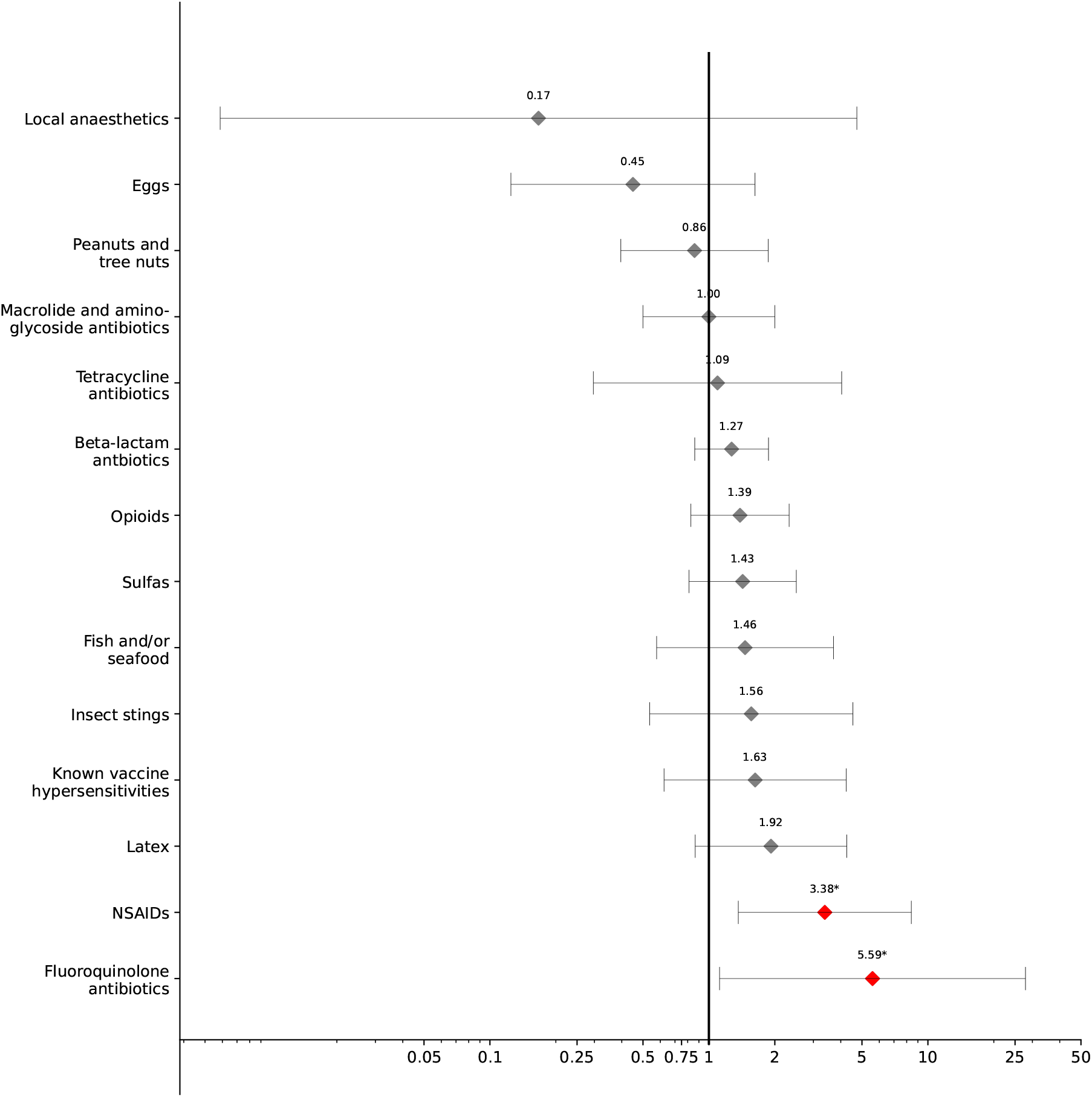
Forest plot of reporting odds ratio of anaphylactic reactions upon receipt of an mRNA vaccine by reported prior allergic status.

## 4 Discussion

### 4.1 mRNA vaccines are not associated with a higher risk of anaphylaxis

We hypothesised that mRNA-based vaccines do not have a specific anaphylactogenic property, i.e. that they are not associated with a statistically significant elevated odds ratio in the whole population. The overall ROR after matching for age and gender of 1.325 (95% CI: 1.212 – 1.448), significant at *p* < 0.001, confirms this hypothesis.

Age and gender have a clear differential effect on the likelihood of developing an anaphylactic reaction following mRNA vaccination. However, the size of this effect is extremely modest. In both cases, the highest RORs with statistical significance are still below 2.0, indicative of a weak association but no safety signal of note.

The conclusions of this study lend further support to the evidence that mRNA-based vaccines are not unusually anaphylactogenic in all age groups and genders. This contribution goes beyond COVID-19 vaccines, and may serve as a building block of a wider case for the safety of mRNA-based vaccinations. There is no evidence of a specific anaphylactogenic effect above and beyond other vaccines when it comes to mRNA-based vaccines. This finding may help in addressing legitimate fears of anaphylaxis both in clinical practice and in the wider communication of public health measures.

### 4.2 Previous allergies do not predispose to higher risk of anaphylaxis from mRNA vaccines

From the analysis above, it appears that no previous allergy significantly predisposes to anaphylaxis following mRNA vaccination, even if in a population that has previous allergies, anaphylactic reactions might, unsurprisingly, be marginally more frequent. Two statistically significant data points stand out from the rest. Previously known allergies to NSAIDs (ROR: 3.382, 95% CI: 1.362 – 8.399) and fluoroquinolone antibiotics (ROR: 5.586, 95% CI: 1.119 – 27.900) might correlate to a somewhat elevated odds ratio of reporting anaphylactic reactions. The source of this effect, as well as its physiological basis, remains open for further research. One might consider, for instance, that NSAID-induced bronchoconstriction is significantly more frequent among patients with asthma,^[19]^ which in turn tends to correlate with higher rates of anaphylaxis. Thus, the presence of a known NSAID allergy might in fact reflect a predisposition to atopy and allergic reactions, and therefore result in higher odds of anaphylaxis. An alternate hypothesis may be that at least some NSAID allergies, such as those to ketorolac (Toradol), might be triggered by the tromethamine ligand present in some formulations (in the case of ketorolac, ketorolac tromethamine), as well as the Moderna vaccine (but not the Pfizer/BioNTech vaccine). Evidence is currently not sufficiently strong to either support or reject such a hypothesis, either. Further research on predisposing factors is clearly needed, and absent more evidence, it cannot be said with confidence that persons with previous allergic histories to NSAIDs or fluoroquinolone antibiotics are at a higher risk of developing anaphylactic adverse effects following mRNA vaccinations. For clinical practice, emphasis should remain on the safety of mRNA vaccination in patients even with extensive prior allergic histories.

The risk from such predispositions must, of course, be considered in the quantitative context. The ROR is at best modestly increased for both NSAIDs and fluoroquinolones, and is overall vanishingly small when considered against the number of vaccinations. Our data record 36 cases of anaphylaxis in individuals with a reported hypersensitivity to fluoroquinolones and 70 cases of anaphylaxis in individuals with a reported hypersensitivity to NSAIDs, out of approx. 328 million vaccination events, corresponding to one event per 9.11 million vaccinations (fluoroquinolones) and one event per 4.85 million vaccinations (NSAIDs). In the absence of a plausible biological causal mechanism, there is no reason to conclude for or against a causal relationship. Given the sparsity of evidence in this field, as well as the risks from COVID-19 in particular for populations who already suffer from respiratory conditions, e.g. inflammatory airway remodeling from asthma, this finding does not justify advising against the vaccine for persons with any known prior allergies.

### 4.3 mRNA vaccines are unaffected by prior vaccine allergies

The relatively novel method of action of mRNA vaccines means that many of the standard allergens that are frequently blamed for post-vaccine anaphylactic and anaphylactoid reactions are absent. The absence of virions means that there is no risk of a hypersensitivity arising from culture media, such as persons with egg allergies tend to experience with yellow fever vaccinations and certain influenzavirus vaccines cultured in eggs. Using a low-temperature cold chain means both mRNA vaccines currently approved under Emergency Use Authorizations for COVID-19 are preservative-free.

The consequence of this novel methodology appears to be reflected by the lack of an association between anaphylactic events from mRNA vaccines and a history of previous hypersensitivity reactions to vaccinations. This study has failed to find a statistically significant association between anaphylaxis and previous vaccination-related hypersensitivity events. While further research and monitoring in this area, too, remains necessary, the absence of such a finding might reassure patients with previous experiences of vaccine-induced anaphylaxis and allay fears of such reactions from mRNA vaccines.

### 4.4 Limitations

This study relied strongly on reporting from VAERS. As all studies that utilise passive reporting data, it is limited by the biases inherent in reporting itself. These biases, by definition, are imperfectly understood.^[20–22]^ Underreporting, in which an adverse effect is not reported, is a frequent confounder, as is overreporting, where the same event is reported multiple times, often due to an innocent lack of communication (e.g. the parent of a vaccinated child makes a report, unsure whether the physician will also make a report). To some extent, underreporting is mitigated for the mRNA-based vaccines currently approved. Both mRNA vaccines are administered under Emergency Use Authorizations that mandate reporting to VAERS of any serious adverse effects, whereas such mandatory reporting is typically only applicable for other vaccines if they fall within the scope *ratione materiae* of the National Childhood Vaccine Injury Act’s mandatory reporting provisions. Since anaphylaxis almost always necessitates treatment, frequently results in hospitalisation and is considered a serious, indeed life-threatening, medical emergency by providers, it is very likely that the rate of underreporting is quite low. Because non-mRNA vaccines, except for the Janssen COVID-19 vaccine, are not held to this standard, it is quite likely that there is a bias from differential underreporting. The likelihood of reporting an anaphylactic or anaphylactoid reaction to VAERS is higher for COVID-19 vaccines, including both mRNA vaccines, than non-COVID-19 vaccines. For this reason, the reporting odds ratios may exaggerate the risk of anaphylactic reactions for mRNA-based vaccines, and thus these odds ratios may be better considered to be rational upper bounds or ‘worst case scenarios’ than actual values.

For previous allergic aetiologies, text mining using regular expressions was utilised. This necessarily could only comprise certain cases, and it is quite certain that due to differential spelling or other issues, some cases were not adequately identified. We tried to compensate for this through regular expressions that are robust to such issues, but the risk of slight biases from differential spelling and phrasing can never be completely excluded when analysing a free-text field.

## 5 Conclusion

Our analysis of anaphylactic events between 01 January 2000 and 02 July 2021 indicates no statistically significant increase in the risk of anaphylactic or anaphylactoid adverse events in the context of mRNA-based vaccines. As such, it adds to a growing body of evidence in favour of the safety and tolerability of such vaccines.^[23–25]^ Of particular note is that neither age nor gender appear to constitute significant, non-trivial differential risk factors for anaphylaxis following mRNA vaccination. Furthermore, previous allergic reactions to vaccines are also not associated with a statistically significant increase in risk. While there is a slight increase in risk for persons with known hypersensitivities to NSAIDs and/or fluoroquinolone antibiotics, the precise extent of this, as well as the underlying physiological mechanism, remains for further study.

This early analysis of anaphylactic adverse events in the context of mRNA vaccinations may be integrated into both clinical practice and the wider field of public health communications. It supports a tentative assertion that mRNA vaccination is not associated with a higher risk of reporting an anaphylactic reaction than non-mRNA vaccination. The safety of this novel and highly effective class of vaccines, and its tolerability specifically in groups who have experienced previous hypersensitivity reactions with vaccines and are thus reluctant to get vaccinated, is becoming the subject of an ever growing body of evidence. This study contributes to that by casting light specifically on the aspect of anaphylactic and anaphylactoid reactions, concluding that the evidence does not raise concerns that could justify non-vaccination.

## Data Availability

VAERS reporting data is available from the CDC's website at https://vaers.hhs.gov.
All code and scripts supporting this manuscript are deposited at
https://github.com/chrisvoncsefalvay/covid-19-vaccine-anaphylaxis and are made available under the DOI 10.5281/zenodo.5112367.

https://vaers.hhs.gov/data.html

https://github.com/chrisvoncsefalvay/covid-19-vaccine-anaphylaxis

## Supplementary material

Supplementary material S1, a CSV (comma-separated values) version of the data underlying Figures 3 and 2, is available as part of DOI 10.5281/zenodo.5112367.

## Funding

This research was funded by Starschema Inc. under its intramural research funding programme.

## Data availability

VAERS reporting data is available from the CDC’s website at https://vaers.hhs.gov. All code and scripts supporting this manuscript are deposited at https://github.com/chrisvoncsefalvay/covid-19-vaccine-anaphylaxis and are made available under the DOI 10.5281/zenodo.5112367.

## Conflicts of interest

CvC is a consultant to a company that may be affected by the research reported in this paper. The funders had no role in the design of the study; in the collection, analysis, or interpretation of data; in the writing of the manuscript, or in the decision to publish the results.

## Notes

### Author Declarations

As this is a purely retrospective study over publicly accessible data on VAERS, it is exempt from IRB approval. For this reason, no approval has been sought or obtained.

